# FLOW CYTOMETRY MULTIPLEXED METHOD FOR THE DETECTION OF NEUTRALIZING HUMAN ANTIBODIES TO THE NATIVE SARS-CoV-2 SPIKE PROTEIN

**DOI:** 10.1101/2020.08.24.20180661

**Authors:** Lydia Horndler, Pilar Delgado, Ivaylo Balabanov, Georgina Cornish, Miguel A. Llamas, Sergio Serrano-Villar, Manuel Fresno, Hisse M. van Santen, Balbino Alarcón

## Abstract

A correct identification of seropositive individuals for the Severe Acute Respiratory Syndrome Coronavirus-2 (SARS-CoV-2) infection is of paramount relevance to assess the degree of protection of a human population to present and future outbreaks of the COVID-19 pandemic. We describe here a sensitive and quantitative flow cytometry method using the cytometer-friendly non-adherent Jurkat T cell line that stably expresses the full-length native spike “S” protein of SARS-CoV-2 and a truncated form of the human EGFR that serves a normalizing role. S protein and huEGFRt coding sequences are separated by a T2A self-cleaving sequence, allowing to accurately quantify the presence of anti-S immunoglobulins by calculating a ratio of the mean fluorescence intensities obtained by double-staining with the sera and a monoclonal antibody specific for EGFR. We show that the method allows to detect immune individuals regardless of the result of other serological tests or even repeated PCR monitoring. It can also be employed to detect neutralizing activity in the sera of individuals. Finally, the method can be used in a multiplexed format to simultaneously measure all anti-S human immunoglobulin isotypes in blood and mucosal fluids including total saliva.

## INTRODUCTION

Severe Acute Respiratory Syndrome Coronavirus-2 (SARS-CoV-2) is the causative agent of the global pandemic COVID-19^1^ Phylogenetic analysis of the full genome classifies SARS-CoV-2 as a Betacoronavirus subgenus Sarbecovirus, lineage B and is related to bat isolates of SARS-CoV and is 79% identical to the SARS virus causing a viral epidemic in 2002 \ Like other coronaviruses, SARS-CoV-2 encodes for 16 non-structural proteins at the 5’ end of the genomic RNA and structural proteins spike (S), envelope (E), membrane (M) and nucleocapsid (N) at the 3’ end ^2^. The spike S protein is responsible for binding to ACE2 in host cells which seems to be the main cellular receptor for the virus ^3^. In native state, the S protein forms homotrimers and is composed of two fragments S1 and S that result from proteolytic cleavage of S upon ACE2 binding ^4^. The S1 fragment contains a central RBD sequence that is the actual ACE2-binding sequence and the target of neutralizing antibodies such as those described to neutralize SARS-CoV-1.

Diagnosis of active infection is currently carried out by PCR amplification of viral RNA comprising fragments of the N, E, S and RdRP genes from biological samples taken from bronchoalveolar lavage (BAL), sputum, nasal swabs, pharyngeal swabs and fibronchoscope brush biopsies, with the highest positive rate resulting from PCR of BAL samples ^5^. Positivity in the PCR test vanishes as the infection is resolved although viral RNA shedding, and PCR positivity, could take place even after there is no further production of infective virions and therefore possibility of transmission.

Unlike PCR-based tests, serological tests are not highly valuable to determine what individual has an active infection with SARS-CoV-2 but are key in epidemiology and Public Health policies since, it can provide an estimate of what segment of a population has been infected with the virus and is likely to have acquired total or partial immunity against a possible resurgences of the pandemics. Herd immunity achieved either by natural infection or as a consequence of vaccination is a goal for all health authorities in the world.

Seropositivity is usually established by detecting the presence of IgG or IgM in the serum of individuals using recombinant fragments of the S or N proteins and tests based on ELISA or lateral flow assays ^5,6^. A disadvantage of those tests is that neutralizing antibodies are not directed against the N protein and that recombinant fragments of S miss the quaternary structure of the S protein trimer, which is the native form of the spike protein in the viral envelope. Therefore, possible neutralizing antibodies directed against the native S trimer could be missed in serological tests based on the expression of recombinant proteins.

Flow cytometry of cells that are transfected with vectors that express the S protein represent an attractive alternative strategy for serological tests. This strategy allows detection of antibodies against the native form of the S protein. Such system has been used by transient transfection of HEK293T human cells to analyze different cohorts of COVID-19 patients *^1^*. We have developed here a flow cytometry serological test using stably transfected Jurkat human hematopoietic cells that co-express the native S protein of SARS-CoV-2 and a truncated form of the human EGFR that serves as a normalizer. The Jurkat-S system is amenable to standardization and can be used for multiplexed detection of human immunoglobulins of all isotypes in a single assay. Finally, we show that the system is superior to ELISA-based methods to detect sera of donors containing neutralizing antibodies.

## RESULTS

We have used a lentiviral vector to express the full spike “S” protein of SARS-CoV2 followed by a truncated human EGFR protein (huEGFRt) linked by a T2A self-cleaving sequence in transduced cells. This system allows to express the two proteins from a monocystronic mRNA. We have produced transducing supernatants to express the construct in the human leukemic cell line Jurkat. Cell surface biotinylation followed by immunoprécipitation with streptavidin-agarose beads and western blot with a mix of sera from recovered COVID-19 patients showed a polypeptide of 150 kD when resolve by SDS-PAGE under reducing conditions (Fig. 1b). Interestingly, larger sizes of the immune-reactive polypeptide were resolved in the absence of reducing agents under non-denaturing conditions (i.e., without boiling; Fig. 1c). This data suggest that the S protein of SARS-CoV-2 could be expressed on Jurkat-S plasma membrane as native trimers. To determine if the S protein expressed on Jurkat-S cells was functional, we analyzed if it could promote the formation of syncitia when Jurkat-S cells were co-incubated with the ACE2+ human hepatocarcinoma cell line HepG2. We labelled Jurkat-S cells with the green dye CFSE and HepG2 with the violet dye Cell Trace Violet. After overnight incubation, we detected a Jurkat-S dose-dependent formation of mixed-cell syncitia that were not detected if the HepG2 were incubated with parental Jurkat cells not expressing the S protein (Fig. 1d). This data indicated that the S protein expressed in Jurkat-S cells has fusogenic activity and therefore that it must be in a native conformation.

**Figure 1.**
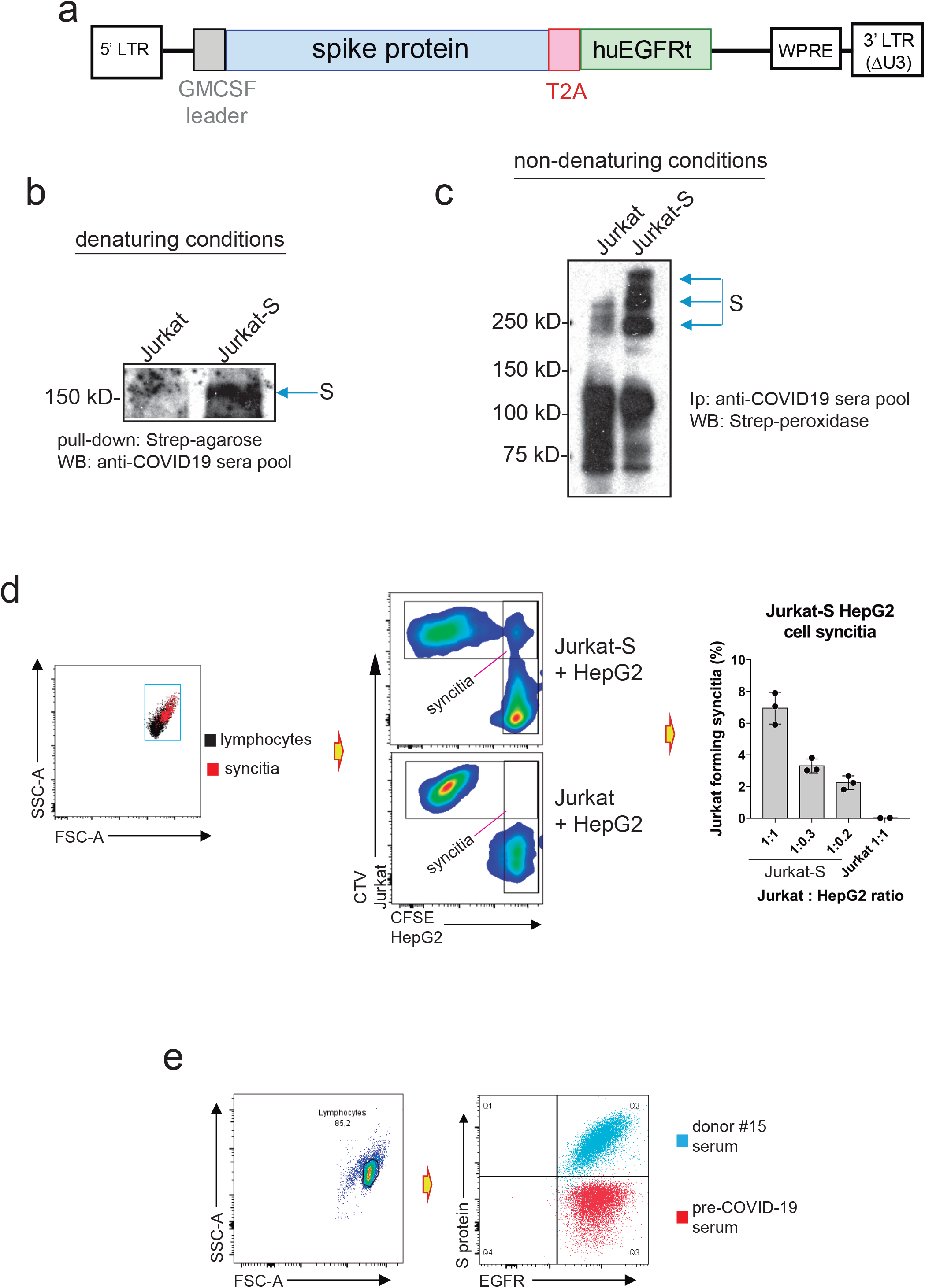
Jurkat-S cells express native SARS-CoV-2 spike S protein allow the detection of anti-S protein antibodies in human sera. **(a)** Lentiviral construct used to permanently express S protein in Jurkat. The full length mature S protein is preceded by a leader sequence of GM-CSF and followed by a T2A sequence which is followed by a tail-less truncated human EGFR construct, **(b)** Expression of S protein on the plasma membrane of Jurkat-S cells assessed by surface biotinylation and followed by pull-down with streptavidin-agarose, SDS-PAGE under reducing conditions and western blot with a mix of sera from donors #15, #3, #4, #8 and #66. The nitrocellulose membrane was finally blotted with a peroxidase-labeled anti-human IgG1 antibody. Biotinylation of the parental Jurkat cells was carried out in parallel as a negative control, **(c)** Expression of the S protein in native form was assessed by surface biotinylation of Jurkat-S and Jurkat cells followed by immunoprecipitation with the mix of sera as above and SDS-PAGE under non-denaturing conditions (i.e. without reducing agents and without boiling). The nitrocellulose membrane was blotted with streptavidin-peroxidase. **(d)** Formation of syncitia between CTV-labelled ACE2+ human cell line and the CFSE-labelled Jurkat-S cells was measured by flow cytometry by analyzing the percentage of cells that become double positive for CTV and CFSE markers. The Bar plot to the right shows the effect of different doses of HepG2 cells on the formation of syncitia with a fixed number of Jurkat. Parental Jurkat cells (not expressing S protein) are considered negative controls. Data represent the mean±SD of triplicated datasets, **(e)** Overlay plot of Jurkat-S cells that were incubated with anti-EGFR mAb conjugated to Bv421 and serum from either donor #15 or from a pre-COVID-19 donor and followed by a secondary anti-human IgG1 antibody conjugated to PE.

The Jurkat-S cells were analyzed using a flow cytometry (FC) assay, where staining with an anti-EGFR monoclonal antibody detects and quatitates expression of the huEGFRt construct alongside detection and quantitation of anti-S protein antibodies within sera from SARS-CoV2-infected blood donors. Figure 1e shows Jurkat-S cells stained with anti-EGFR mAb and either a serum sample taken from an individual before the COVID-19 pandemics (pre-COVID-19 serum) or serum taken from an asymptomatic donor (donor #15) determined positive with multiple serological assays (Table 1). Serum from donor #15 could strongly detect the S protein expressed on Jurkat-S cells whereas pre-COVID-19 serum did not, proving this method to be valid for capture and detection of anti-spike antibodies present in sera from SARS-CoV2-exposed individuals. This assay was highly sensitive and detected anti-S protein antibodies over a wide range of mean fluorescence intensities (MFI) when staining with sera from different blood donors (Figure 2a). In comparison with commercial serological assays this Jurkat-S clone FC assay detected more cases of seropositive individuals (Table 1) and facilitated verification of positive seroconversion in patient samples that had previously given uncertain results (Fig. 2b, orange symbols). Specifically, SARS-CoV2 PCR and serologically negative sera collected from 30 healthcare workers (Hospital Ramón y Cajal, Madrid) in March-May 2020 was re-screened using the Jurkat-S FC assay and detected 2/30 sera (RyC52 a; RyC650) clearly positive for S specific IgG1 (Fig.2c). To increase sensitivity for detecting S specific IgG1 in sera reading MFI values close to the 0.5 threshold for positivity we can plot the EGFR MFI versus S protein MFI (Fig. 1d). S protein expression in Jurkat-S is coupled to EGFRt expression (Fig. 1a), thus positive sera produce a diagonal MFI on a plot of anti-S protein vs EGFR, whereas negative serum generates a flat profile. An example of this can be seen in Fig. 2d with borderline sera RyC58 and RyC70, where RyC58 detects as above borderline positive for S specific IgG1 whilst serum RyC70 remains negative.

**TABLE 1.**
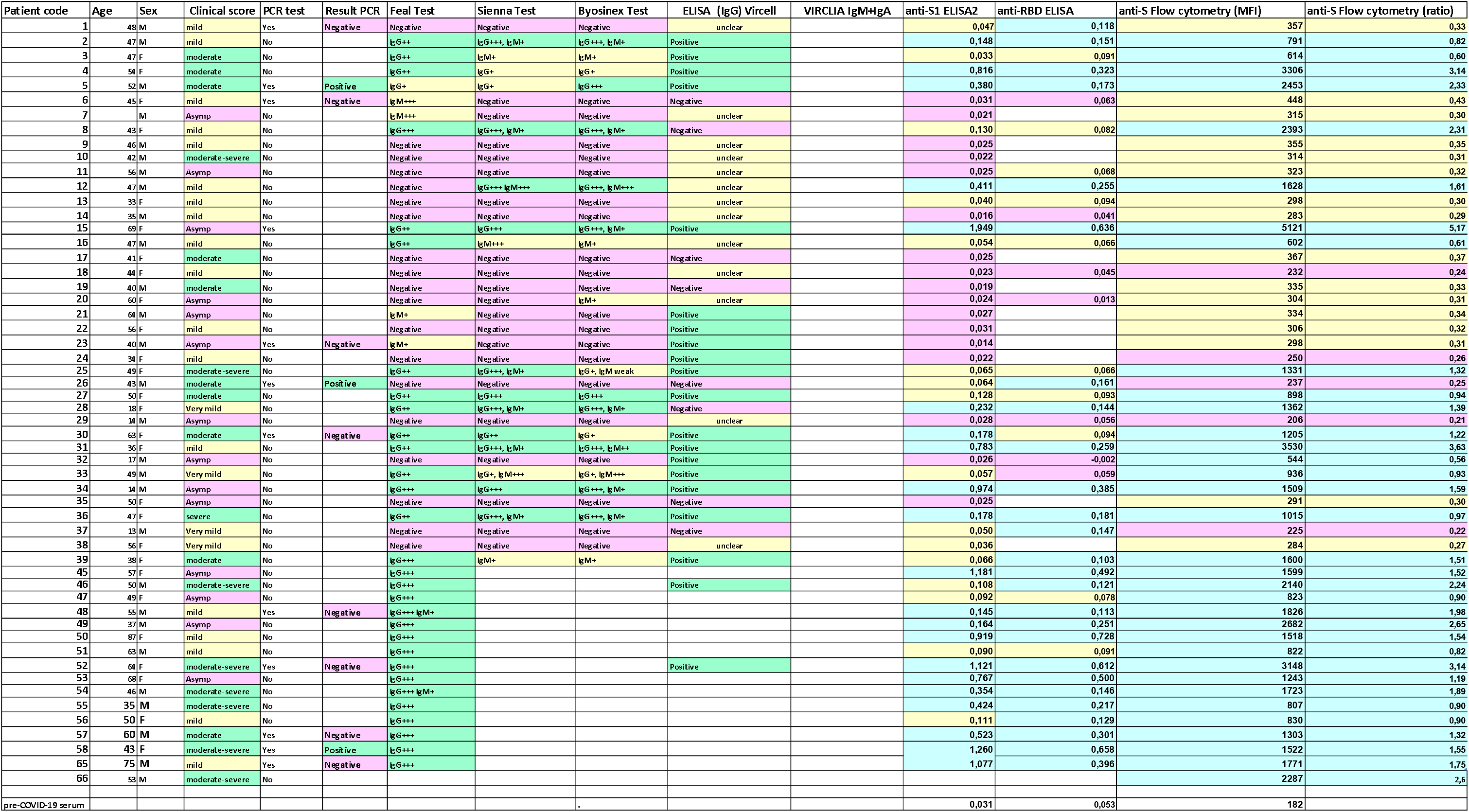
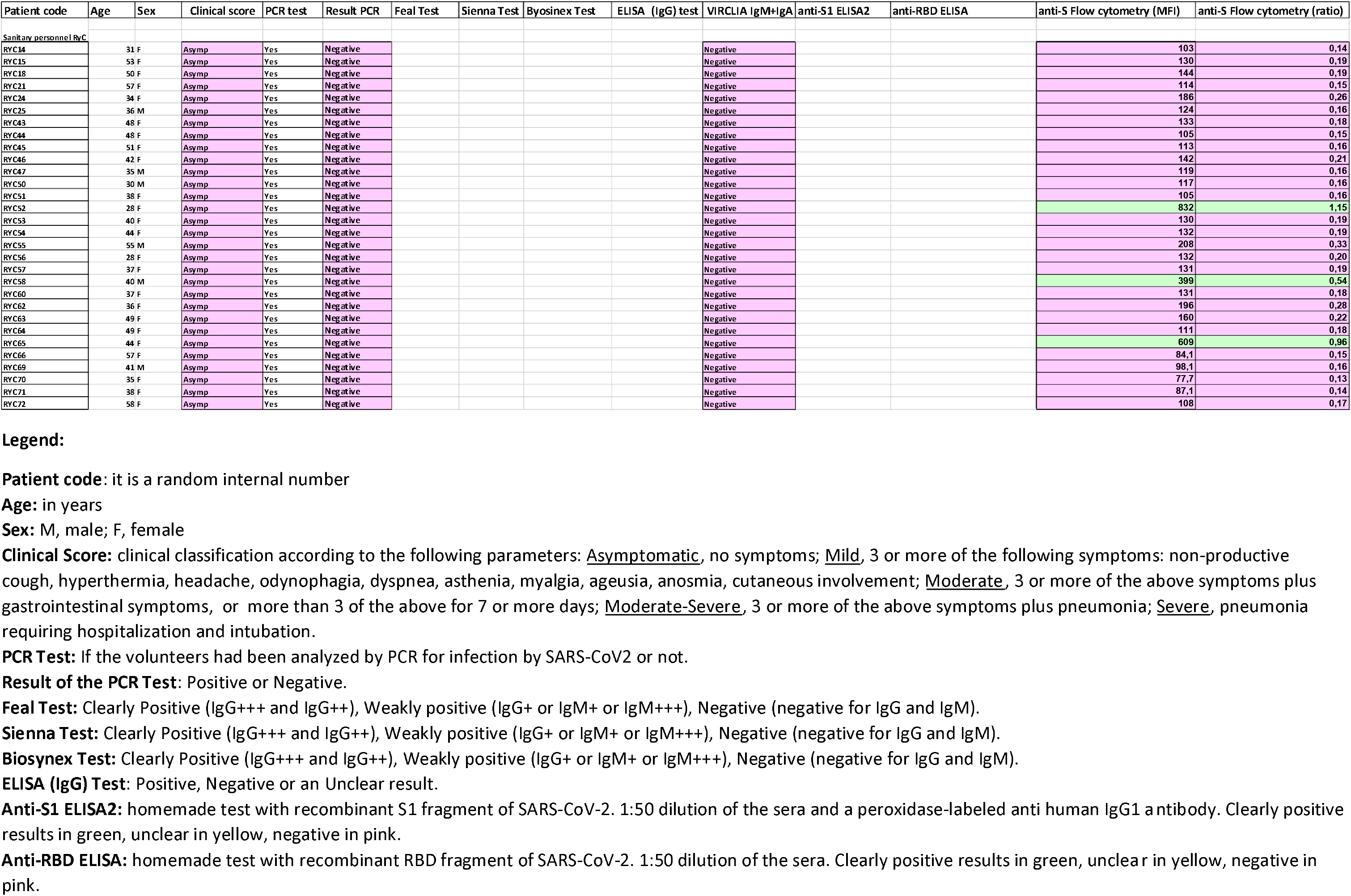
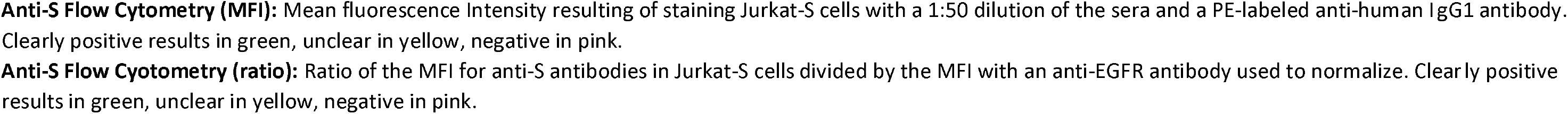

**Figure 2.**
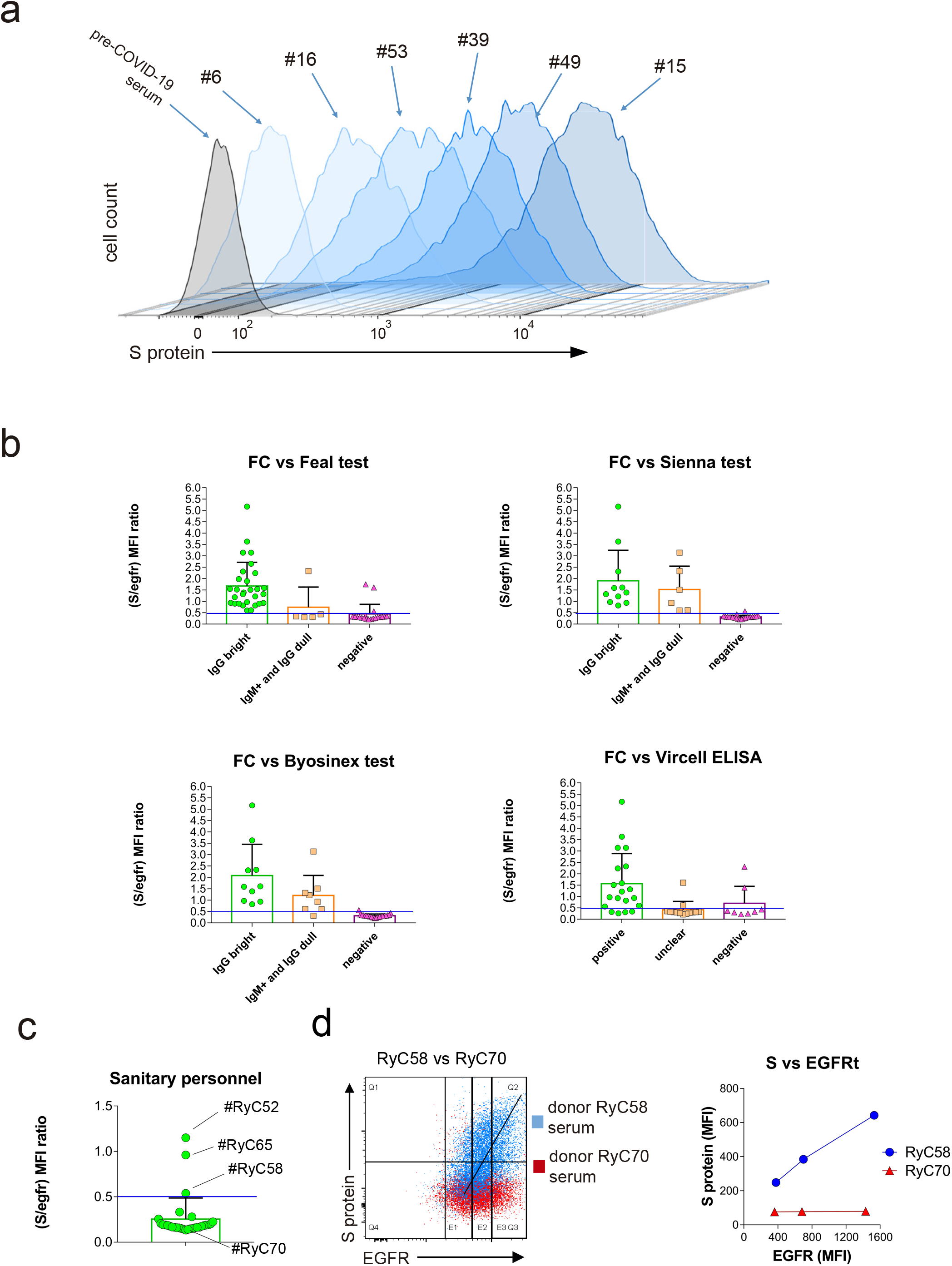
Flow cytometry of Jurkat-S cells allows to detect anti-S immunoglobulins poorly detected by commercial tests with a wide dynamic range, **(a)** Overlay histograms of Jurkat-S staining with different human sera diluted 1:50. A pre-COVID19 serum sample is taken as negative control (grey histogram), **(b)** Bar plots of flow cytometry data generated with Jurkat-S cells and the panel of serum samples of Table 1 classified according to their result in the indicated commercial tests: green, positive for the test; orange, weak or unclear; magenta, negative samples. Flow cytometry data is expressed as the ratio between the MFI of the antibody anti-S and the MFI for EGFR. Negative values for the flow cytometry test are those with a S/EGFR MFI ratio lower than 0.5. **(c)** Bar plot showing the S/EGFR MFI ratio determined by flow cytometry analysis with Jurkat-S of serum samples from 30 sanitary personnel repeatedly tested as PCR negative for SARS-CoV-2 at the Ramón y Cajal Hospital of Madrid. A negative result is considered for a ratio lower than 0.5. Two clear cases of sera positive for anti-S IgG1 are indicated (RyC52 and RyC65). A borderline sample just above the threshold line (RyC58) is also indicated, **(d)** Two-color dot plot of anti-S protein fluorescence versus EGFR fluorescence for borderline serum sample RyC58 (blue) and the clearly negative RyC70 sample (red). The line plot to the right shows the MFI for EGFR taken at the 3 sectors indicated in the two-color plot and the corresponding MFI values for EGFR.

In order to compare the sensitivity of our Jurkat-S assay with a conventional ELISA, we received a kind gift of recombinant Spike proteins, specifically the S1 and receptor binding domain (RBD) fragments, from Dr. Peter Cherepanov (CRICK, London). We coated the plates with 2 mg/ml of either S1 or RBD and incubated the coated plates with a 1:50 dilution of human sera. Binding of human IgG1 antibodies was detected using a peroxidase-labeled anti-human IgG1 monoclonal antibody. Comparing absorbance values from the ELISA with MFI values of the Jurkat-S FC assay across sera stratified by experienced COVID-19 symptoms (asymptomatic, mild, moderate, moderate-severe and severe) showed the MFI values to be spread across a wider range of values than abosorbance values (Fig. 3a). Finally, the comparison of Absorbance values in the two ELISA tests (anti-S1 and anti-RBD) produced a good-fitted straight line, whereas the comparison of the FC MFI with the absorbance values (against S1 and RBD) does not adjust to a straight line (Figure 3b). It is clear that detecting S-specific IgG1 using the Jurkat-S FC assay increases sensitivity for detecting SARS-CoV2-exposure in individuals testing negative by ELISA. These individuals may have generated antibodies against other fragments of the Spike, e.g. S2^8^, or against the native trimeric structure of the S protein, that are found on the surface of Jrukat-S cells but not in the ELISAs. Sera from donors #8, #46, #48 and #49 were apparently negative for anti-S IgG1 by ELISA but clearly positive by FC with Jurkat-S. By contrast, anti-S IgG1 was detected in sera from donors #15, #31 and #52 by both ELISA and FC, and serum from donor #58, appeared to be detected better by ELISA than by FC (Fig. 3b). To determine if those differences were maintained at different dilutions of the sera, a titration test was carried out in parallel using the FC Jurkat-S method and ELISA of the S1 fragment. The results showed that all sera, including that of donor #58, were clearly positive by FC even at a 1:450 dilution whereas by ELISA, sera #8, #46, #48 and #49 remained negative (Fig. 3c). These results indicate that sera, which could have been considered negative by ELISA, are indeed positive for S-specific IgG1 by ELISA. Sample #49 was from an asymptomatic individual but sera #8, #46 and #48 were from individuals who had experienced symptoms that ranged from mild to severe (Table 1). Next we assayed sera testing positive by FC and negative by ELISA for the capacity to neutralize S protein function. We generated pseudotyped lentiviral reporter particles coated either with the S protein of SARS-CoV2 or with the irrelevant G protein of VSV virus, as a control. The viral particles were used to transduce HEK293T cells stably transfected with ACE2 (Fig. 4a). Transduction of the ACE2+ HEK293T cells was inhibited in a dose-dependent manner by sera from donors #66 and #15 that were identified as positive by both the ELISA and FC methods (Fig. 3c and Table 1) (Fig. 4a). The effect of the #66 serum was due to specifically neutralizing the S protein since this serum did not inhibit transduction of ACE2+ HEK293T cells by pseudotyped VSV G protein lentivirus (Fig. 4a). Interestingly, serum from donor #48 and, to a lesser extent, from donors #8 and #49, were also able to neutralize the S protein pseudotyped lentivirus (Fig. 4a), suggesting that these serum samples contain neutralizing antibodies despite being seronegative by ELISA (Fig. 3b and 3c). These data show that the Jurkat-S FC assay can be superior to ELISA for detecting protective immunity to SARS-CoV2.

**Figure 3.**
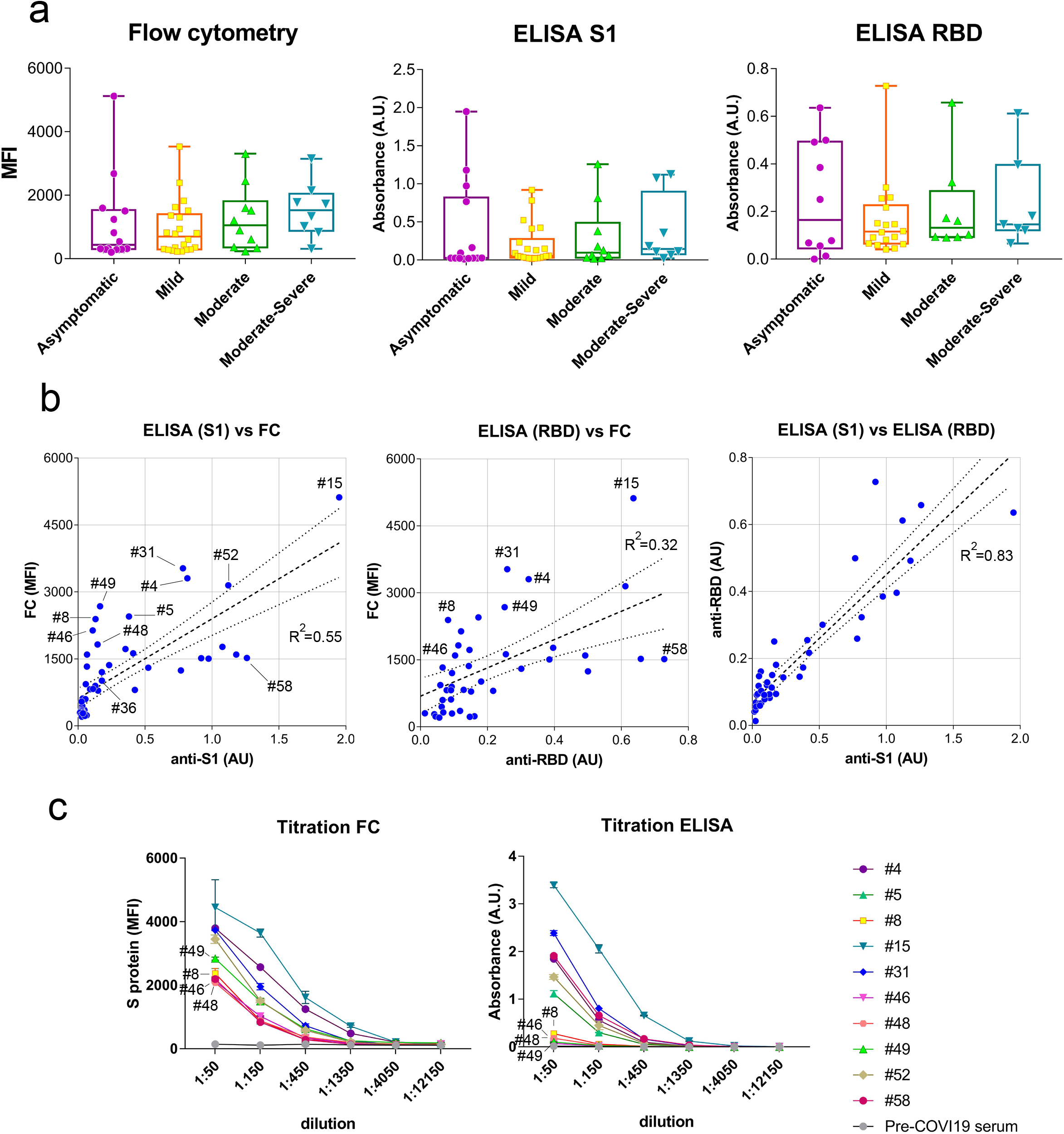
Flow cytometry of Jurkat-S cells detects anti-S antibodies in sera otherwise negative for ELISA using recombinant S1 and RBD proteins, **(a)** Box and whiskers plot of MFI and absorbance data in human sera according to their clinical classification of COVID-19 symptoms. Clinical classification was done according to the following parameters: Asymptomatic, no symptoms; <u>Mild</u>, 3 or more of the following symptoms: non-productive cough, hyperthermia, headache, odynophagia, dyspnea, asthenia, myalgia, ageusia, anosmia, cutaneous involvement; <u>Moderate</u>, 3 or more of the above symptoms plus gastrointestinal symptoms, or more than 3 of the above for 7 or more days; <u>Moderate-Severe</u>, 3 or more of the above symptoms plus pneumonia; <u>Severe</u>, pneumonia requiring hospitalization and intubation, **(b)** Comparison of MFI versus absorbance data generated by flow cytometry and ELISA for all human sera. A lineal regression curve was adjusted, with a 95% confidence interval, to all data with the R^2^ values indicated in the plots. Selected samples of outliers (#4, #8, #31, #49) as well as samples close to a diagonal (#36, #12, #52, #15) are Indicated, (c) Titration of selected human sera by ELISA using the S1 protein and by flow cytometry with Jurkat-S cells. Samples with antibodies detected by flow cytometry and not by ELISA are indicated (#8, #46, #48, #49). Data represent the mean±SD of triplicates.

We moved onto multiplex the Jurkat-S FC assay for the detection of a panel of anti-S immunoglobulins, including IgG1, IgG2, IgG3, IgG4, IgA and IgM. We detected all anti-S isotypes in several serum samples and calculated the S/EGFR MFI ratio for each (Table 2). The most robust binding was observed for IgG1 in all cases, followed by far by IgG4 and IgA (Fig. 4b). A weak IgM response, in terms of S/egfr MFI ratio, was detected in all samples with the highest value in sample RyC65, suggesting that this asymptomatic health worker may have only been recently infected when bled. The presence of anti-S antibodies of the IgG2 and IgG3 subclasses was not clearly detected in all samples. Figure 4c shows a summary heatmap of the fold change in detection of each anti-S immunoglobulin isotype, referred to the MFI of the pre-COVID-19 sample. This indicates that the predominant humoral response to the S protein in serum is in the form of IgG1 followed by IgM and IgG4.

**TABLE 2.**

|  | MFI<br>IgG1 | MFI<br>egfr | MFI<br>IgG3 | MFI<br>IgG4 | MFI<br>IgA | MFI<br>IgM | MFI<br>IgG2 | ratio<br>IgG1/egfr | ratio<br>IgG3/egfr | ratio<br>IgG4/egfr | ratio<br>IgA/egfr | ratio<br>IgM/egfr | ratio<br>IgG2/egfr | ratio<br>IgG2 |
| --- | --- | --- | --- | --- | --- | --- | --- | --- | --- | --- | --- | --- | --- | --- |
| #8 | 3555 | 876 | 809 | 748 | 596 | 140 | 716 | 4,058219178 | 0,923515982 | 0,853881279 | 0,680365297 | 0,159817352 | 0,817351598 | 0,8173516 |
|  | 3691 | 892 | 892 | 765 | 715 | 154 | 846 | 4,137892377 | 1 | 0,857623318 | 0,801569507 | 0,17264574 | 0,948430493 | 0,94843049 |
|  | 3540 | 817 | 849 | 685 | 669 | 130 | 819 | 4,332925337 | 1,039167687 | 0,838433293 | 0,818849449 | 0,159118727 | 1,00244798 | 1,00244798 |
| #48 | 1502 | 854 | 634 | 139 | 555 | 192 | 671 | 1,758782201 | 0,742388759 | 0,162763466 | 0,649882904 | 0,224824356 | 0,785714286 | 0,78571429 |
|  | 1607 | 882 | 805 | 181 | 726 | 214 | 839 | 1,821995465 | 0,912698413 | 0,20521542 | 0,823129252 | 0,242630385 | 0,951247166 | 0,95124717 |
|  | 1580 | 901 | 762 | 166 | 639 | 195 | 793 | 1,753607103 | 0,84572697 | 0,184239734 | 0,709211987 | 0,216426193 | 0,880133185 | 0,88013319 |
| #49 | 2087 | 916 | 819 | 323 | 726 | 262 | 875 | 2,278384279 | 0,894104803 | 0,352620087 | 0,792576419 | 0,286026201 | 0,955240175 | 0,95524017 |
|  | 2096 | 919 | 877 | 342 | 796 | 309 | 918 | 2,280739935 | 0,95429815 | 0,372143634 | 0,866158868 | 0,336235038 | 0,998911861 | 0,99891186 |
|  | 2044 | 915 | 923 | 326 | 842 | 302 | 989 | 2,233879781 | 1,008743169 | 0,356284153 | 0,920218579 | 0,330054645 | 1,080874317 | 1,08087432 |
| #66 | 2287 | 863 | 825 | 446 | 605 | 280 | 880 | 2,650057937 | 0,955967555 | 0,516801854 | 0,701042874 | 0,324449594 | 1,019698725 | 1,01969873 |
|  | 2243 | 870 | 892 | 439 | 667 | 270 | 935 | 2,57816092 | 1,025287356 | 0,504597701 | 0,766666667 | 0,310344828 | 1,074712644 | 1,07471264 |
|  | 2156 | 831 | 785 | 426 | 569 | 260 | 819 | 2,594464501 | 0,944645006 | 0,512635379 | 0,684717208 | 0,312876053 | 0,985559567 | 0,98555957 |
| #15 | 2108 | 953 | 792 | 211 | 708 | 342 | 798 | 2,211962225 | 0,831059811 | 0,221406086 | 0,742917104 | 0,358866737 | 0,837355719 | 0,83735572 |
|  | 2194 | 953 | 1122 | 277 | 946 | 430 | 1069 | 2,302203568 | 1,177334732 | 0,29066107 | 0,992654774 | 0,451206716 | 1,121720881 | 1,12172088 |
|  | 1941 | 899 | 715 | 188 | 642 | 287 | 692 | 2,159065628 | 0,795328142 | 0,209121246 | 0,714126808 | 0,319243604 | 0,76974416 | 0,76974416 |
| RyC58 | 749 | 791 | 443 | 96,2 | 348 | 128 | 494 | 0,946902655 | 0,560050569 | 0,121618205 | 0,439949431 | 0,16182048 | 0,624525917 | 0,62452592 |
|  | 726 | 730 | 459 | 101 | 365 | 124 | 487 | 0,994520548 | 0,628767123 | 0,138356164 | 0,5 | 0,169863014 | 0,667123288 | 0,66712329 |
|  | 768 | 726 | 500 | 110 | 409 | 136 | 534 | 1,05785124 | 0,688705234 | 0,151515152 | 0,563360882 | 0,187327824 | 0,73553719 | 0,73553719 |
| RyC65 | 982 | 777 | 627 | 119 | 458 | 429 | 556 | 1,263835264 | 0,806949807 | 0,153153153 | 0,589446589 | 0,552123552 | 0,715572716 | 0,71557272 |
|  | 948 | 765 | 635 | 152 | 506 | 494 | 582 | 1,239215686 | 0,830065359 | 0,19869281 | 0,661437908 | 0,645751634 | 0,760784314 | 0,76078431 |
|  | 945 | 728 | 524 | 102 | 466 | 437 | 521 | 1,298076923 | 0,71978022 | 0,14010989 | 0,64010989 | 0,600274725 | 0,715659341 | 0,71565934 |
| RyC70 | 739 | 751 | 474 | 107 | 380 | 156 | 519 | 0,984021305 | 0,631158455 | 0,142476698 | 0,505992011 | 0,207723036 | 0,691078562 | 0,69107856 |
|  | 673 | 734 | 432 | 87,5 | 360 | 166 | 476 | 0,916893733 | 0,588555858 | 0,119209809 | 0,490463215 | 0,226158038 | 0,648501362 | 0,64850136 |
|  | 729 | 762 | 499 | 105 | 422 | 201 | 543 | 0,956692913 | 0,654855643 | 0,137795276 | 0,553805774 | 0,263779528 | 0,712598425 | 0,71259843 |
| pre-CoVID19 | 702 | 728 | 479 | 139 | 366 | 57,5 | 478 | 0,964285714 | 0,657967033 | 0,190934066 | 0,502747253 | 0,078983516 | 0,656593407 | 0,65659341 |
|  | 729 | 752 | 544 | 166 | 368 | 59,3 | 537 | 0,969414894 | 0,723404255 | 0,220744681 | 0,489361702 | 0,078856383 | 0,714095745 | 0,71409574 |
|  | 777 | 720 | 563 | 179 | 421 | 81,8 | 588 | 1,079166667 | 0,781944444 | 0,248611111 | 0,584722222 | 0,113611111 | 0,816666667 | 0,81666667 |

**Figure 4.**
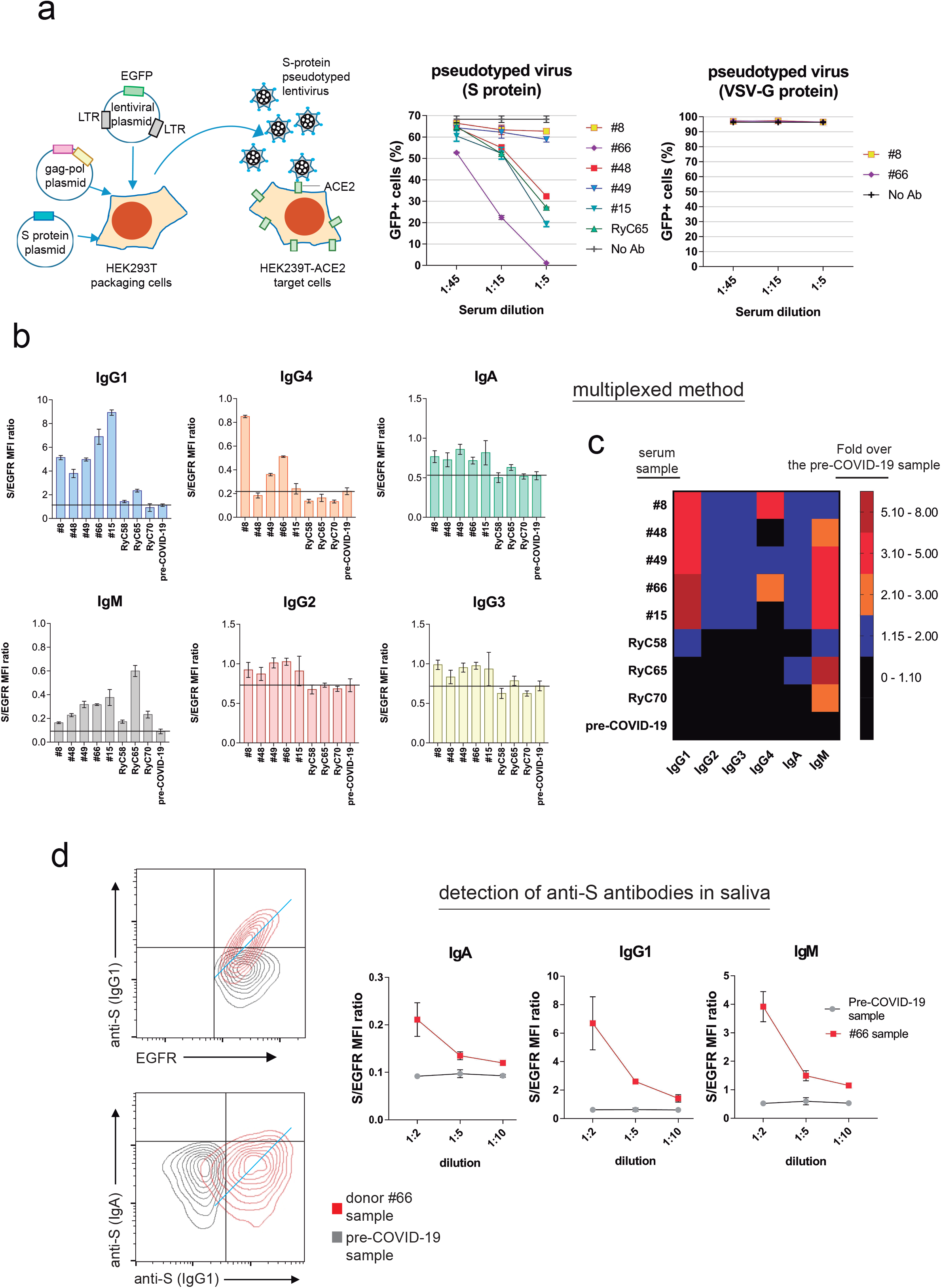
Flow cytometry of Jurkat-S cells can be used in multiplexed methods to detect simultaneously the expression of anti-S antibodies of all subclasses with neutralizing activity, **(a)** Neutralization of infection with pseudo-typed lentivirus by sera from COVID-19 patients. Cartoon of the strategy for generation of pseudotyped lentiviruses (left). Neutralization of lentiviruses pseudotyped with either the S protein (center) or the VSV G protein (right) by the indicated dilutions of human sera. Data represent the mean±SD of triplicates, **(b)** Detection of anti-S immunoglobulins of the indicated subclasses in a multiplexed method of flow cytometry in the serum samples indicated at 1:50 dilution. Data represent the mean±SD of triplicated datasets, (c) Rainbow heatmap plot of the anti-S protein antibody data shown in b, normalized as fold value of the S/egfr MFI ratio in each sample divided by the mean S/egfr MFI ratio of the pre-COVID-19 serum, **(d)** Detection of anti-S antibodies of **IgA** but also of IgG1 and IgM isotypes in total saliva from a seropositive donor. Two-color plots are shown to illustrate the direct relationship between anti-S IgG1 binding and EGFR binding. Also to show IgA versus IgG1 binding to Jurkat-S. Data represent the mean±SD of triplicates.

The FC method on Jurkat-S cells was also used to determine the presence of anti-S immunoglobulins in total saliva of donor #66 compared with the content in a sample from a seronegative donor. The method allowed the detection of IgA but also IgG1 and IgM in the saliva sample of donor #66 in a dose-dependent fashion and above the concentrations detected in a seronegative control sample (Fig. 4d).

## DISCUSSION

Here we describe a method based on flow cytometry of a hematopoietic cell line that stably expresses the S protein of SARS-CoV-2 in its native form. Compared to methods based on the use of recombinant purified proteins, the FC method with Jurkat-S offers a wider range of response and it is highly quantitative. The coordinated expression of huEGFRt marker serves the function of normalizing marker and of unambiguously determining the positivity of sera with low antibody titers. The system can also be used in a multiplex format to simultaneously detect all immunoglobulin subclasses specific for the S protein and to detect the possible presence of neutralizing antibodies in mucosal fluids of easy access such as saliva. Importantly, it is superior to current ELISA methods to determine the fraction of the human population that has acquired antibody responses and neutralizing activity towards SARS-CoV2.

A FC-based method has been previously described using HEK293T cells that overexpress the S protein^7 8^. Compared to this, the Jurkat-S system described here offers the advantage of employing a non-adherent cell line that does not require methods such as trypsinization to place them in suspension. In addition, as said above, the coordinated expression of the huEGFRt marker from a single polypeptide makes it very convenient to resolve ambiguous samples. This property has served to detect as seropositive 3 cases of a cohort of 30 sanitary personnel highly exposed to SARS-CoV-2 and repeatedly considered as negative by PCR and other serological methods.

Compared to ELISA and rapid serological tests, the FC Jurkat-S method has the disadvantage of requiring the use of a flow cytometer. However, the extended use of those machines in the immunology and hematology departments of hospitals may make the method of choice if the goal is to accurately determine what proportion of a human population likely has a protective humoral response and how far is from reaching the anxiously sought herd immunity. In summary, we believe that this method may represent an excellent addition to COVID-19 Epidemiology and Public Health policies.

## MATERIALS AND METHODS

### Cells

The human T-cell line Jurkat clone E6-1 was acquired from ATCC (TIB-152) were maintained in complete RPMI 1640 supplemented with 5% fetal bovine serum (FBS, Sigma) in a 5% CO2 incubator. Human embryonic kidney HEK293T cells (ATCC CRL-3216) and human hepatocellular carcinoma HepG2 cells (ATCC HB-8065) were maintained in DMEM supplemented with 10% FBS in a CO2 incubator. HEK293T cells expressing ACE2 were generated by lentiviral transduction with vector CSIB and selection in blasticidin S ^8^ All cell lines were routinely tested for the absence of mycoplasma.

### Lentiviral vector and Jurkat cell transduction

To express the full-length spike S protein of SARS-CoV2 we used the lentiviral vector based on the epHIV-7 plasmid that contains the truncated version of human EGFR (huEGFRt) that lacks the domains essential for ligand binding and tyrosine kinase activity described in Wang et al ^9^. For transduction, lentiviral-transducing supernatants were produced from transfected packaging HEK-293T cells as described previously ^10^. Briefly, lentiviruses were obtained by co-transfecting plasmids pCMV-dR (gag/pol) and using the JetPEI transfection reagent (Polyplus Transfection). Viral supernatants were obtained after 24 and 48 hours of transfection. Polybrene (8 μg/mL) was added to the viral supernatants prior to transduction of Jurkat cells. A total of 3×10^5^ Jurkat cells were plated in a P24 flat-bottom well in 350 μL of DMEM and 350 μL of viral supernatant were added. Cells were centrifuged for 90 minutes at 2200 rpm and 32^9^C and left in culture for 24 hours. Transduced cells were selected by FACS sorting 48 h later using the anti-EGFR antibody.

### Human sera

A total of 84 human sera were obtained from volunteers that contacted EMPIREO SL (www.empireo.es) for serological tests. Each participant provided written consent to participate in the study which was performed according to the EU guidelines and the Declaration of Helsinki. Serum donors filled in a questionnaire to allow their clinical classification according to the following parameters: <u>Asymptomatic</u>, no symptoms; <u>Mild</u>, 3 or more of the following symptoms: non-productive cough, hyperthermia, headache, odynophagia, dyspnea, asthenia, myalgia, ageusia, anosmia, cutaneous involvement; <u>Moderate</u>. 3 or more of the above symptoms plus gastrointestinal symptoms, or more than 3 of the above for 7 or more days; <u>Moderate-Severe</u>. 3 or more of the above symptoms plus pneumonia; <u>Severe</u>, pneumonia requiring hospitalization and intubation. A second set of 30 serum samples was obtained from health care workers at the Intensive Care Unit of the Ramón y Cajal Hospital in Madrid. Those workers have been routinely tested for COVID-19 by PCR and resulted always negative.

### Flow cytometry

Jurkat-S cells were incubated for 30 min on ice with 1:50 dilutions of human sera in phosphate-buffered saline (PBS), 1% bovine serum albumin (BSA), 0.02% sodium azide. Cells were spun for 5 min at 900 g and the pellet was resuspended in PBS-BSA buffer and spun to eliminate the excess of antibody. Two additional washes were carried out. The cell pellet was finally resuspended in a 1:200 dilution of mouse anti-human IgG1 Fc-PE (Ref.: 9054-09, Southern Biotech) and a 1:300 dilution of the Brilliant Violet 421™ anti-human EGFR Antibody (Ref.: 352911, Biolegend) in PBS-BSA. Samples were then washed and labeled cells were analyzed on a FACSCalibur or FACSCanto II flow cytometer (Becton-Dickinson) and the data were analyzed with FlowJo software (BD). For multiplexing, the following antibodies were used (all from Cytognos, S.L.): FITC-labelled anti-IgG1, PE-labeled anti-IgG2, APC-labelled anti-IgG3, APCC750 labelled anti-IgG4, PEcy7-labelled anti-IgM andPerCPcy5.5-labelled anti-IgA (for IgA1 and IgA2).

### Commercial kits

The following commercial serological tests were assayed:

-Feal Test of Hangzhou Alltest Biotech, reference number RPP25COV1925.

-Sienna Test of Salofa Oy, reference number 102221.

-BiosSynex Test of Biosynex Swiss, reference number SW40005.

-Vircell ELISA test of Vircell SL, reference number G1032.

-COVID-19 VIRCLIA IgM+IgA monotest, Vircell SL, reference number VCM098.

### ELISA

96 well plates (MaxiSorp NUNC-Immunoplate) were coated overnight at 4 °C with S1 (2 μg/ml) and were subsequently blocked for 1 h with 1% BSA (Sigma). Coated plates were incubated with the diluted sera for 1.5 h at room temperature. Bound antibodies were detected by incubation with mouse anti-human IgG1 secondary antibody coupled to horse-radish-peroxidase (HRP) (Southern Biotech) diluted 1/6,000 in 1% BSA in PBS which was then detected using an ABTS substrate solution (Invitrogen). The OD at 415 nm was determined on a iMark microplate reader (Bio-Rad). Serum from a healthy individual previous to COVID-19 was used as negative control.

### Immunoprecipitation and Western Blot

For surface biotinylation, 20 × 10^6^ Jurkat-S cells were incubated for 45 min on ice with 0.5 mg/ml of sulfo-NHS-biotin (Pierce) in PBS supplemented with 0.1 mM CaCl_2_ and 1 mM MgCl_2_. After washing, the cells were lysed in Brij96 lysis buffer (0.33% Brij96, 150 mM NaCl; 20 mM Tris-HCl pH 7.8; 10 mM iodoacetamide; 1 mM PMSF; 1 μg/ml aprotinin; 1 μg/ml leupeptin) and pull-down was carried out Streptavidin-agarose beads. The bead-bound material was washed 5 times and subjected to SDS-PAGE. The proteins recovered were then transferred to a nitrocelullose membrane that was probed with with a mix of sera from donors #15, #3, #4, #8 and #66, diluted 1:2000. The nitrocellulose membrane was developed by ECL (Pierce) using PO-coupled mouse anti-human secondary antibodies (Southern Biotech).

### Neutralization assay with pseudotyped virus

Lentiviral supernatants were produced from transfected HEK-293T cells as described previously (Martinez-Martin et al., 2009). Briefly, lentiviruses were obtained by co-transfecting plasmids psPAX2 (gag/pol), pHRSIN-GFP and either a truncated S envelope (pCR3.1-St)^11^ or VSV envelope (pMD2.G) using the JetPEI transfection reagent (Polyplus Transfection). Viral supernatants were obtained after 24 and 48 hours of transfection. Polybrene (8 μg/mL) was added to the viral supernatants prior to transduction of ACE2+HEK293T cells. A total of 3×10^5^ MOLT-4 cells were plated on a P24 flat-bottom well 350 μL of DMEM and 350 μL of viral supernatant were added. Cells were centrifuged for 90 minutes at 2200 rpm and left in culture for 24 hours.

### Formation of syncitia

To analyze the formation of syncitia between Jurkat-S and HepG2 cells, HepG2 cells were detached from the culture plate by trypsinization and resuspended at a concentration of 3×106 cells/ml in PBS. Jurkat-S cells were collected by centrifugation and resuspended in PBS at the same concentration. HepG2 cells were labelled with Cell Trace Violet (CTV, Invitrogen) for 5 min at 37°C in PBS; Jurkat-S were labelled with CFDA-SE (CFSE, Invitrogen) under the same conditions. Both dyes were used at a final concentration of 5 μM. Labelling was stopped by adding complete medium and the cells washed with medium and finally mixed at different ratios in complete RPMI medium + 5% FBS and plated overnight at 37<u>^o^</u>C. Formation of syncitia was analyzed by flow cytometry by calculating the percentage of cells double positive for CFSE and CTV.

## Data Availability

All data is contained within the results of the manuscript and is available to the reader

## Acknowledgments

We are indebted to Valentina Blanco and Tania Gómez for their expert technical assistance. We thank Dr. Peter Cherepanov, Annachiara Rosa and Chloe Roustan (Crick COVID-19 Consortium, Francis Crick Institute, London, UK) for a generous gift of recombinant SARS CoV2 S1 and RBD antigens. We also thank Dr. P. Beniasz, The Rockefelloer University, for providing constructs.

## Author contributions

LH performed research and analyzed the data; PD and IB helped with ELISAs and other experimentation; GC provided the original idea and edited the manuscript, MALL and SS-V provided clinical samples and data, MF and HMvS analyzed data and supervised research, BA supervised and designed research, analyzed data and wrote the manuscript.

## REFERENCES

1. Lu, R. et al. Genomic characterisation and epidemiology of 2019 novel coronavirus: implications for virus origins and receptor binding. The Lancet 395, 565–574 (2020).

2. Su, S. et al. Epidemiology, Genetic Recombination, and Pathogenesis of Coronaviruses. Trends Microbiol. 24, 490–502 (2016).

3. Hoffmann, M. et al. SARS-CoV-2 Cell Entry Depends on ACE2 and TMPRSS2 and Is Blocked by a Clinically Proven Protease Inhibitor. Cell 181, 271–280.e8 (2020).

4. Walls, A. C. et al. Structure, Function, and Antigenicity of the SARS-CoV-2 Spike Glycoprotein. Cell 181, 281–292.e6 (2020).

5. Venter, M. & Richter, K. Towards effective diagnostic assays for COVID-19: a review. J. Clin. Pathol. 73, 370–377 (2020).

6. Weissleder, R., Lee, H., Ko, J. & Pittet, M. J. COVID-19 diagnostics in context. Sci. Transi. Med. 12, eabcl931 (2020).

7. Grzelak, L. et al. SARS-CoV-2 serological analysis of COVID-19 hospitalized patients, pauci-symptomatic individuals and blood donors. http://medrxiv.org/lookup/doi/10.1101/2020.04.21.20068858 (2020) doi: 10.1101/2020.04.21.20068858.

8. Ng, K. W. et al. *Pre-existing and* de novo *humoral immunity to SARS-CoV-2 in humans.* http://biorxiv.org/lookup/doi/10.1101/2020.05.14.095414 (2020) doi: 10.1101/2020.05.14.095414.

9. Wang, X. et al. A transgene-encoded cell surface polypeptide for selection, in vivo tracking, and ablation of engineered cells. Blood 118,1255–1263 (2011).

10. Martinez-Martin, N. et al. Cooperativity between T cell receptor complexes revealed by conformational mutants of CD3epsilon. Sci Signal 2, ra43. (2009).

11. Robbiani, D. F. et al. Convergent antibody responses to SARS-CoV-2 in convalescent individuals. Nature (2020) doi:10.1038/s41586-020-2456-9.

